# Forecasting trends of HIV infection using deep learning models in East Gojjam zone, North West Ethiopia, 2025

**DOI:** 10.1101/2025.10.05.25337384

**Authors:** Daniel shitu, Andualem fentahun Senishaw, Zegeye Regasa Hordofa, Maru meseret Tadele, Gizaw Hailiye Teferi

## Abstract

**Background:** The growing burden of HIV/AIDS, particularly in sub-Saharan Africa, presents a significant public health challenge, characterized by increasing morbidity, and mortality rates. This region is disproportionately affected, bearing for two-thirds of the global HIV/AIDS problem, highlighting an urgent need for effective solutions. Accurate forecasting of new HIV infections is crucial for developing targeted interventions to combat the HIV/AIDS pandemic.

**Objective:** This study aims to forecast trends of new HIV infections for the next five years and identify the contributing factors in the East Gojjam Zone.

**Methods:** DHIS2 (2018-2025) data set from East Gojjam zone were analyzed using to a hybrid machine learning and deep learning framework. Machine learning models (Decision Tree, Random Forest, XGBoost, LightGBM, CatBoost, AdaBoost, and Gradient Boosting) were used for feature selection, and deep learning architectures (RNN, LSTM, GRU, and bidirectional variants) were used for time-series forecasting. Model performance was assessed using MAE, MSE, RMSE and MAPE

**Result:** From the seven machine-learning algorithms used for selecting important futures the random forest was best performed model and many features were selected to apply for further forecasting using deep learning algorithms. Bidirectional LSTM model was best performed model among the six sequential deep learning algorithms used for forecasting HIV infection in East Gojjam zone. Forecasts reveal an upward trend of HIV infection in study area.

**Conclusion:** Combination of Machine learning and Deep learning algorithms method shows high predictive accuracy in forecasting of HIV infection. The forecasted trend shows an upward trend and needs urgent intervention and attention to combat the problem.

## Background

Human Immune Virus (HIV) is a retrovirus that weakens the immune system by attacking CD4 cells, potentially leading to AIDS, which increases vulnerability to infections and cancers. It spreads mainly through unprotected sex, sharing contaminated needles, and from mother to child during pregnancy, birth, or breastfeeding [2, 3].

HIV/AIDS remains a major global health challenge, especially in Sub-Saharan Africa, which accounts for about 70% of cases [3]. Since the epidemic began, around 85.6 million people have been infected and 40.4 million have died. By the end of 2022, 39 million people were living with HIV, with global adult prevalence at 0.7%, and 3.2% in the WHO African Region [4].

Forecasting HIV/AIDS is vital for managing comorbidities, drug resistance, and rising mortality. In Ethiopia, the traditional AIM model is used, but it lacks data-driven learning and has several limitations [5, 6]. Recently, there has been growing interest in using artificial intelligence in healthcare. Machine learning, especially deep learning, shows strong potential for analysing large medical datasets and overcoming limitations of traditional methods. These advanced algorithms can make accurate, data-driven predictions, making them valuable tools for forecasting HIV/AIDS trends and guiding timely interventions [7, 8].

Forecasting new HIV infections is crucial for efficient resource use, treatment planning, and prevention. It enables early intervention, improves outcomes, and supports on-going program evaluation. Using deep learning algorithms enhances prediction accuracy, helping health systems respond more effectively to the HIV/AIDS burden [9–13].

Despite on-going care and monitoring, HIV/AIDS remains a significant public health issue, with rising cases in some high-transmission areas…[14]. The disease poses a major global challenge, especially in Sub-Saharan Africa, which accounts for two-thirds of all cases [15]. In Ethiopia, an estimated 19,999 HIV-related deaths were reported, with studies showing high mortality rates—45% linked to neurological issues at Tikur Ambesa Hospital, and 6.7 and 5.15 deaths per 100 person-years at Metema and in the Somalia region, respectively[16, 17] [18].

Several studies have examined HIV trends worldwide. Global new infections peaked at 3.16 million in 1999, dropping to 1.94 million by 2017 [19]. In Ghana, ARIMA-based forecasts showed rising cases, especially among those over 30 [20]. Similarly, a study in the Philippines employing deep learning techniques projected cumulative HIV cases to reach 145,273 by 2030 [20]. In Kazakhstan, an ARIMA model forecasted an increase in HIV prevalence from 0.29 in 2021 to 0.47 by 2030[21]. In China, neural network algorithms indicated a continued increase in HIV incidence from May 2018 to December 2020[22]. Additionally, an analysis based on the Age-Period-Cohort model highlighted an upward trend in HIV incidence among individuals aged 41 to 70 years from 2005 to 2015 [23]. Furthermore, machine learning algorithms, particularly random forest, demonstrated high performance in predicting HIV infection among men who have sex with men in China[24]. These studies collectively provide insights into HIV trends and the efficacy of various forecasting and prediction methodologies.

In the Amhara region of Ethiopia, the new HIV infection rate was reported at 6.63 per 1,000 tested adults from 2015 to 2017, with a total of 35,210 new cases [25]. From July 2019 to August 2021, the Amhara Public Health Institute reported that 14.2% of new infections occurred in this region from among the HIV positive clients tested by Asante HIV recent test[26]. The rising incidence of HIV/AIDS causes serious health challenges, including higher morbidity, treatment failures, healthcare costs, and mortality—requiring urgent action. While few forecasting studies exist globally, none have been done in Ethiopia’s East Gojjam zone. This gap limits effective response, making this study vital for informing policymakers and guiding future strategies.

By leveraging machine learning, we can develop more adaptive and precise forecasting models, ultimately improving our strategies and responses to the dynamic landscape of HIV/AIDS[5, 6].

### Factors associated with HIV infection (incidence) from previous literatures

There are many risks for HIV infections, those are being the young age [27], being female sex [28], adults who had primary, secondary and higher educational levels had higher odds of being HIV positive than non-educated individuals [29], in contrast to this other study revealed lower educational status has highest HIV risk as compared to higher educational status [25], rural residence as a risk for HIV[30], migrants has higher risk[25], rural residence of HEI [31] higher in Nyanza province as compared to Nairobi [32], similarly higher in geographic region north central Uganda as compared to central Uganda/Kampala [28], increased wealth index has risk for HIV infection [29, 32], current marital status of widowed women has high HIV risk as compared to never married women [28] and also a divorced women as compared to never married women [28], similarly in other study showed that widowed women has high risk HIV infection as compared to non-polygamous married women [32] and also a divorced women as compared to non-polygamous married women[32], women who were 1 of 3 or more wives [32], history of modern medical injection[27], invasive instrumentation [27], Multi-partner sex [27–29, 33], men having sex with men[34], commercial sex workers[34], STIs[27, 33], HSV-2 infection[28, 33], [27], history of illegal abortion[27], risky sexual behaviour[27], Not using condoms with partners outside of marriage as compared to not having outside partner[28], uncircumcised men [28, 32], home delivery [30, 35, 36], infant not receiving ARV prophylaxis at birth [30, 36], mixed feeding practices [30, 35–37] and mother-child pairs neither receiving ARV therapy [30, 37], mothers being on late AIDS stage [35], absence of maternal PMTCT interventions[35, 38], episiotomy[36], HIV exposed infants mothers without ANC[31], less than 350 CD4 count of HIV exposed infants mothers [31], shorter duration of HIV treatment increase the risk of mother-to-child transmission of HIV[39], Injection drug users [40], tattoos [40], history of imprisonment [40], genetic factors like protective effect of CCR5 D32 variant, CCR5HHE carrying *59402A is associated with increased likelihood of infection and development of AIDS, low CCL3L1 copy numbers are related to increased susceptibility to HIV infection[41], occupational exposures risk factors for HIV infection like Contact to potentially infectious body fluids, needle stick injury and glove breakage [42].

This study aims to identify important features for HIV infection, model development for forecasting HIV infection, and forecasting the trends of HIV infection for next five years in East Gojjam zone

## Methods

### Study area and period

The study was conducted in the East Gojjam Zone of Ethiopia’s Amhara Region, with Debre Markos as its capital. Located 265 km from Bahir Dar and 300 km from Addis Ababa, the zone had a population of 2,153,937 according to the 2007 census. As of the 2014 E.C. report, the zone has 11 public hospitals (including one referral, one general, and nine primary), 102 health centers, and 420 health posts.

The study used secondary data from the DHIS2 system, analyzing records from January 1, 2018, to February 1, 2025. It focused on HIV-related services, including PMTCT, TB-HIV, HIV testing, and ART, applying time series analysis to forecast future HIV trends

#### Study design

The study used a retrospective time series design with deep learning algorithms to forecast future HIV infections in East Gojjam. Tailored for time series forecasting, these models captured temporal dependencies and nonlinear patterns in historical data, revealing trends and variations to support accurate, objective-aligned predictions.

### Data source

#### Description of the dataset

The DHIS2 dataset from East Gojjam zone is the study’s primary data source, encompassing health data from health posts, clinics, health centres, and hospitals. While it covers various diseases, only HIV-related data were used.

### Study population and Sample size

This study includes all participants tested for HIV and taking ART care including PMTCT services, TB-HIV co-infection, HIV testing service and People living with HIV (PLWH) in ART care service for forecasting trends of HIV. So the study population for forecasting HIV infection is HIV tested individuals. Sample varies from month to month due to time series nature of data and being aggregate data. Total aggregated study participants for HIV were 1,225,442.

### Inclusion and Exclusion criteria

All patients recorded in DHIS2 under PMTCT, HIV testing, TB-HIV co-infection, and ART care from January 2018 to February 2025 were included. Due to the aggregate nature of the time series data (with dates as rows), no individuals were excluded directly; however, less relevant or sparse features were removed during pre-processing

### Data collection

Data from the DHIS2 system is extracted by DHIS2 trained experts in the East Gojjam zone health office. This comprehensive dataset includes major clinical outcomes such as treatment regimen in different age category, viral load, mortality, nutritional status, family planning status, and other relevant health indicators in ART dataset. In PMTCT dataset includes maternal and child related health information. In TB-HIV confection also includes major health outcomes of HIV and TB treatment related factors. HIV testing service dataset is another which includes major risk groups for HIV like commercial sex workers, long distance drivers, daily laborers and others. Since all HIV care services like ART care is after testing service that is linked after being HIV positive. So those all data sets were extracted from the DHIS2 software in each year starting from 2018 to 2025. Indicators that do not available in currently in webpage were extracted by communicating experts that have national data access mandates at national level that can access previous data versions from the server.

### Pre-processing

Before modelling, extensive pre-processing was performed: datasets were integrated, multi-year data was compiled, and dates were standardized to the Gregorian calendar. Missing values were negligible due to aggregation, but rare zero-value features (e.g., third treatment failure in specific age groups) were removed if >50% sparse or domain-irrelevant. Features were normalized (0–1 scaling) for equal contribution.

During preprocessing of data to model fitting of data different python packages were used namely time converter, pandas, Numpy, Matplotlib, Seaborn, Scikit-learn, keras, tensorflow,Statsmodels, datetime and other libraries were applied. Some of the functions of those libraries are described as follows NumPy serve as a fundamental package for numerical computing, providing support for arrays and mathematical functions[43]. For data reading, managing, and descriptive analysis Pandas was applied [44]. Matplotlib and Seaborn were used for data visualization due its interactive visualizations to analyze data distributions and relationships [45].Scikit-learn were used during model selection, and evaluation. This library includes tools for train test split and many others likes imputation of missing values [46]. For the deep learning aspect of the analysis, Tensor Flow were used as the primary library, providing tools for building and training neural networks[47], and Keras, which runs on top of Tensor Flow used to simplify the process of constructing and training these models[48]. Statsmodels used for estimating and interpreting statistical models, particularly useful in time series analysis like seasonal decomposition.

### Study variables

#### Outcome variable (target)

The new HIV infections target feature of the study

### Independent variables (Features)

All variables available in the DHIS2 in HIV testing service, PMTCT, TB–HIV confection, and ART care were input for the feature selection process and only potential features were used for model development. Machine learning Techniques were employed to identify and retain the most relevant features, ultimately improving the model’s forecasting performance. In above datasets totally two hundred twenty five (225) features in category of HIV testing service related factors, PMTCT related factors, and TB-HIV related factors. After evaluation of 225 variables and removal of empty columns, One hundred fifty six variable(156) remains, then removing unrelated features and features with more than half of zero values or empty, 88 variables were included in feature selection process.

#### Terms and Operational Definitions

Risky sexual behavior is s defined as the occurrence of any of the following activities: sex with commercial sex workers, multiple sexual partners, inconsistent condoms use or failure to use condoms during sexual intercourse with an irregular sex partner and sexual debut at age < 18 years[49]

### Modeling framework

**Figure 1:**
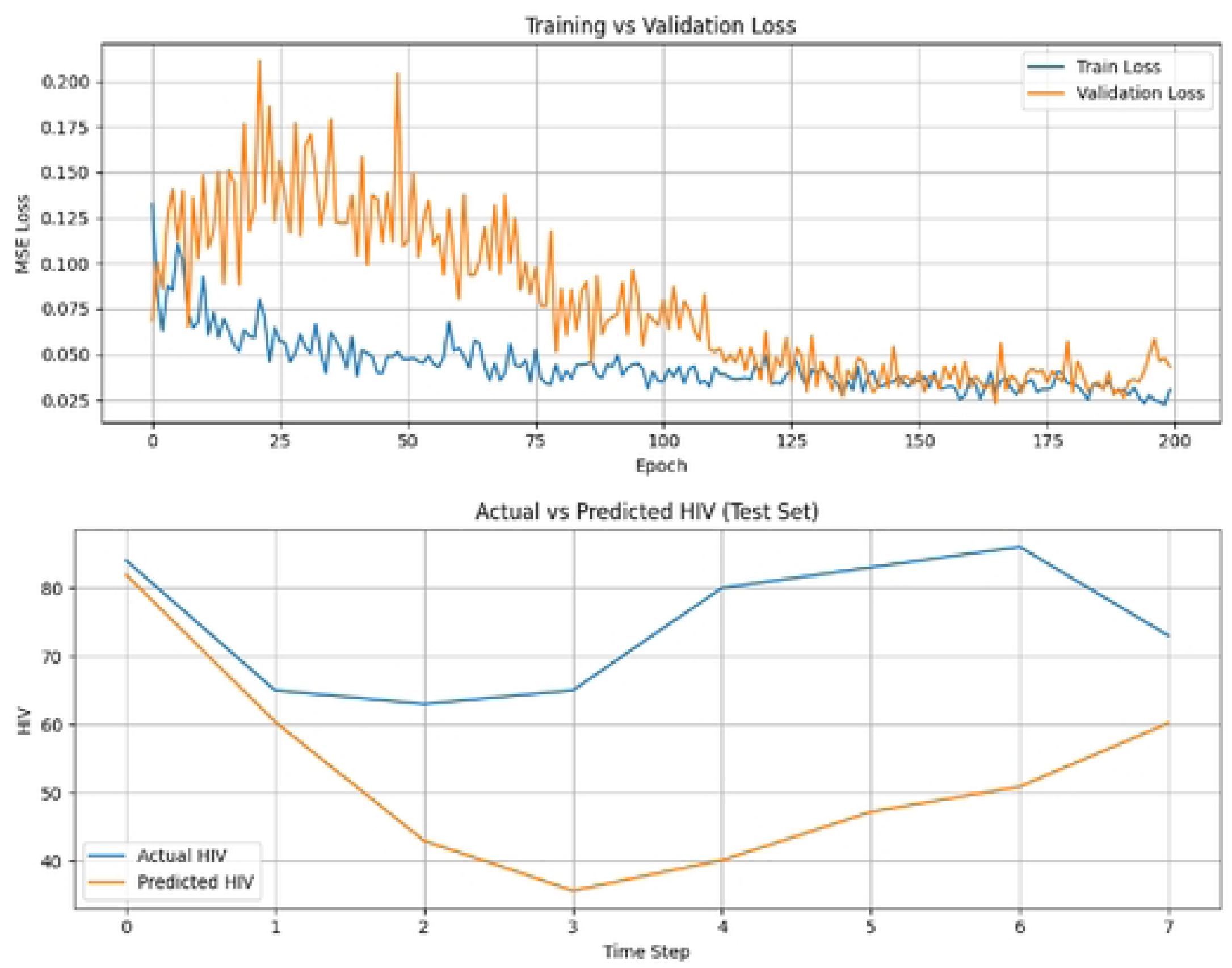
Modeling framework for forecasting HIV infections in East Gojjam zone for the next five years.

#### Models

##### Feature selection methods

This study used a hybrid approach combining machine learning for feature selection and deep learning for time series forecasting. Machine learning models—such as Decision Trees, Random Forest, GBM, XGBoost, AdaBoost, LightGBM, and CatBoost—were applied to identify key factors influencing HIV infection. The best-performing model was selected based on evaluation metrics to support accurate forecasting

##### Model building for forecasting new HIV Infection

This study utilized six deep learning architectures to forecast outcomes over five years. Experiments tested models with up to four layers, ultimately using three-layer structures with dropout and dense layers. ReLU for hidden layers and linear for the outputs performed best. with Adam optimizer used throughout. Hyper-parameters such as learning rate, batch size, epochs, time steps, and dropout were tuned—manual tuning outperformed Grid and Random Search. Training converged best at 200 epochs with an initial batch size of 8. Over-fitting was controlled using dropout (0.6) and early stopping. MSE was used as the loss function, suitable for the regression task.

##### Hyper parameter tuning

The hyper parameter tuning process involved extensive experimentation to identify optimal model configurations. The learning rate was tested at 0.01, 0.001, and 0.0001, with 0.001 selected as the final value. Batch sizes ranging from 1 to 30 were evaluated, settling on 8. For time steps (window size), values from 1 to 60 were tested, with 12 chosen initially, though the model was later retrained using 60 for forecasting. The number of epochs varied between 50 and 200, with 200 proving most effective. Dropout rates between 0.2 and 0.7 were assessed, with 0.6 yielding the best results. For activation functions, ReLU was selected for hidden layers and linear for the output. The optimal architecture consisted of 3 layers with neuron configurations of (110, 70, 50), chosen after testing various arrangements. The Adam optimizer and Mean Squared Error [50] loss function were employed throughout. Early stopping and lag variables (up to six) were tested but did not improve performance and were excluded from the final model. The data was split into training, validation, and test sets (80%, 10%, 10%), which performed better than the 70-15-15 split. Manual hyperparameter tuning proved most effective compared to random and quick search approaches.

#### Model evaluation methods

Evaluating time series deep learning models like RNNs, LSTMs, GRUs, and their bidirectional variants involves both quantitative metrics and visual analysis. Key metrics include MAE, MSE, RMSE, and MAPE—capturing average errors, emphasizing larger ones, translating errors into target units, and providing scale-independent comparisons. These metrics are essential for assessing model accuracy and reliability [51–53]. Model evaluation methods for machine learning algorithms also used mean squared error for feature selection for HIV infection and first line treatment failure.

In addition to numerical metrics, visual evaluation methods are crucial for interpreting model performance[54]. Prediction plots visually compare predicted and actual values, revealing how well the model captures data patterns. Combined with quantitative metrics, they help ensure robust real-world performance.

#### Ethical clearance

Ethical clearance was obtained from Debre Markos University’s Health Sciences Ethical Review Committee, and then approved by the Amhara Public Health Institute and East Gojjam Zone Health Office. Permission was granted to access DHIS2 data from all East Gojjam zone health facilities. As secondary data were used, client confidentiality was protected through data anonemization.

#### Result

##### Overview of HIV infection East Gojjam zone

Monthly Trends of new HIV infections in East Gojjam Zone

**Figure 2:**
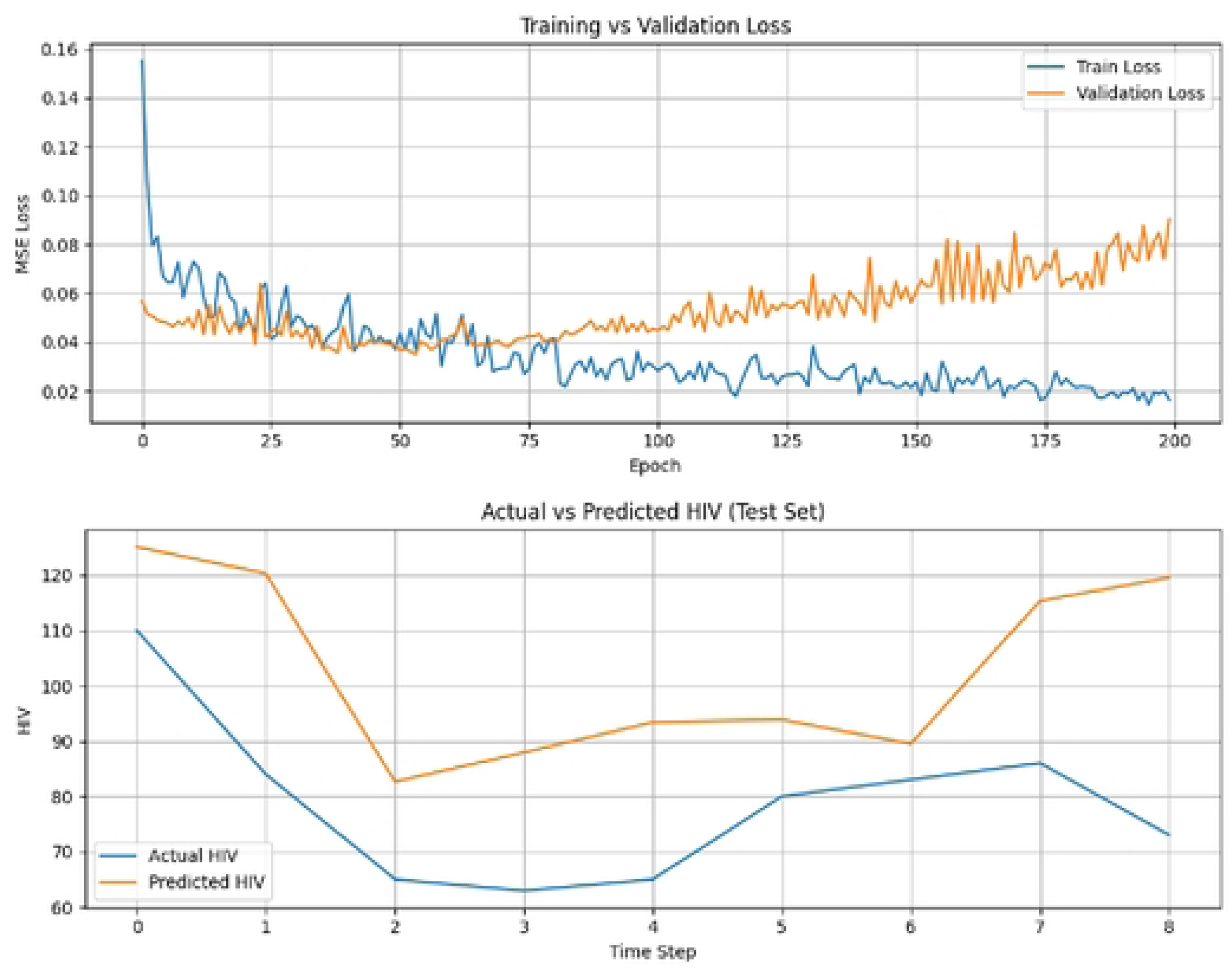
trends of new HIV infections in East Gojjam zone.

**Figure 3:**
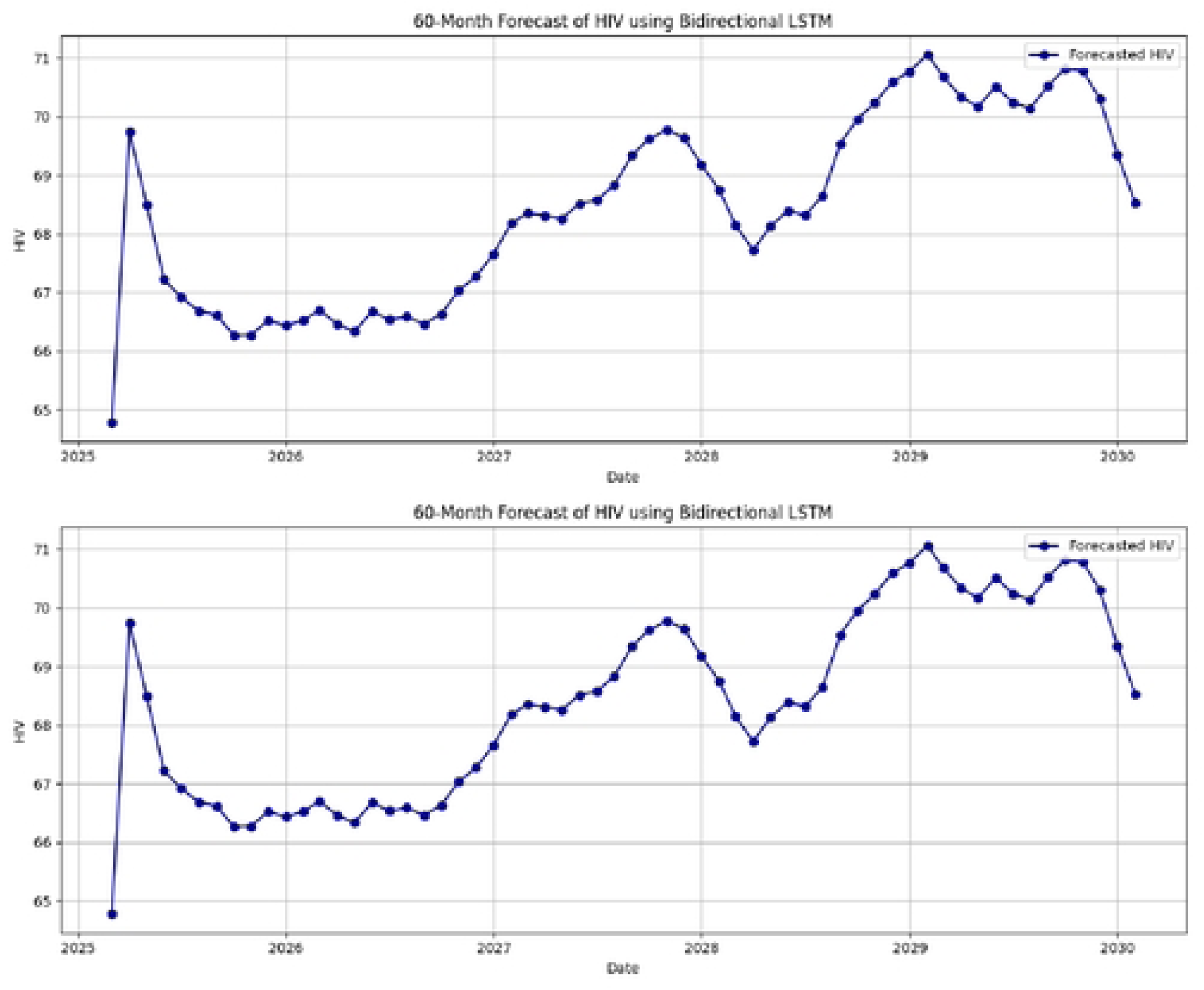
Seasonal decomposition of HIV infection.

Two outlier observations were found in the data set and could have negatively influenced model training and forecasting performance by introducing noise and bias figure 1. The outliers were replaced with values close to neighbouring data points to preserve the continuity of the time series and trend, and limited the risk of over fitting or instability in the forecasting models.

**Figure 4:**
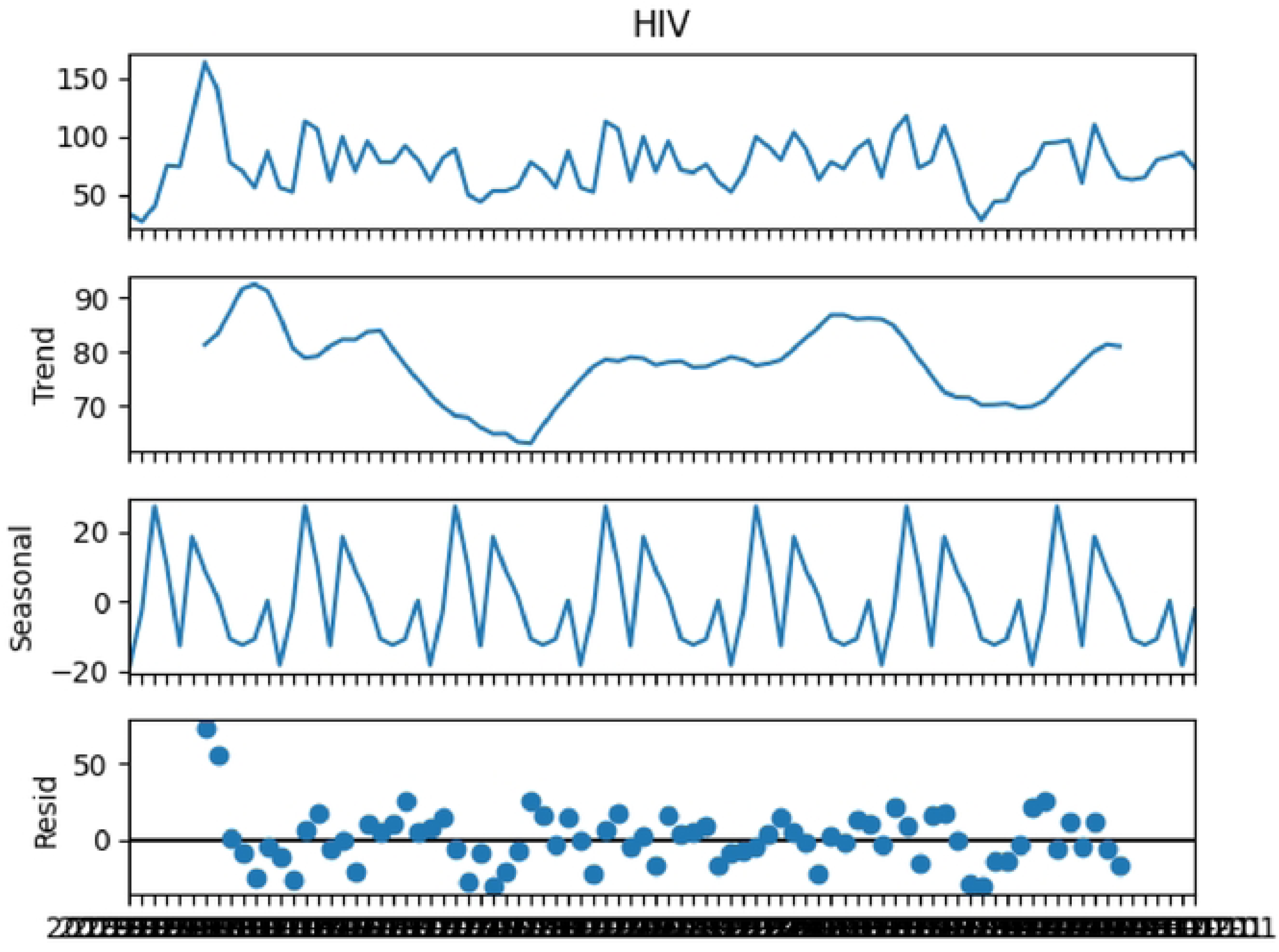
outlier of HIV infection in East Gojjam zone.

#### Important feature selection for HIV infection

**Table 1:** Machine earning algorithms for feature selection for HIV infection.

Based on the mean square error value the random forest score was the lowest among the rest of above machine learning algorithms. So that the features selected by random forest were used to forecast future five years of new HIV infection magnitude in the East Gojjam zone.

**Figure 5:**
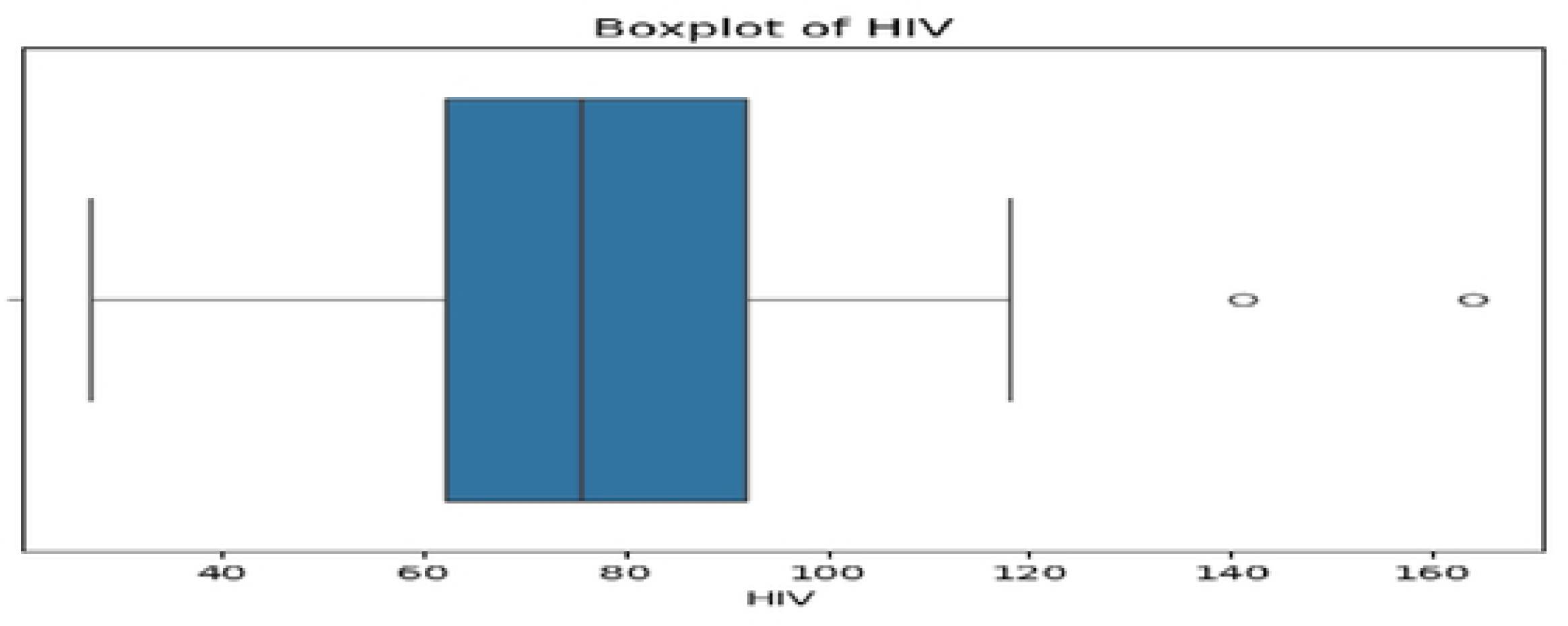
important features selected for HIV using random forest.

#### HIV forecasting using time series deep algorithms

**Table 2:** Comparisons of deep learning model performance in forecasting HIV infection in East Gojjam zone using quantitative evaluation metrics.

##### RNN for Forecasting HIV infection in East Gojjam zone

Test MAE (original scale): 8.1801 Test MSE: 102.5954

Test RMSE: 10.1289

Test MAPE: 11.27%

**Figure 6:**
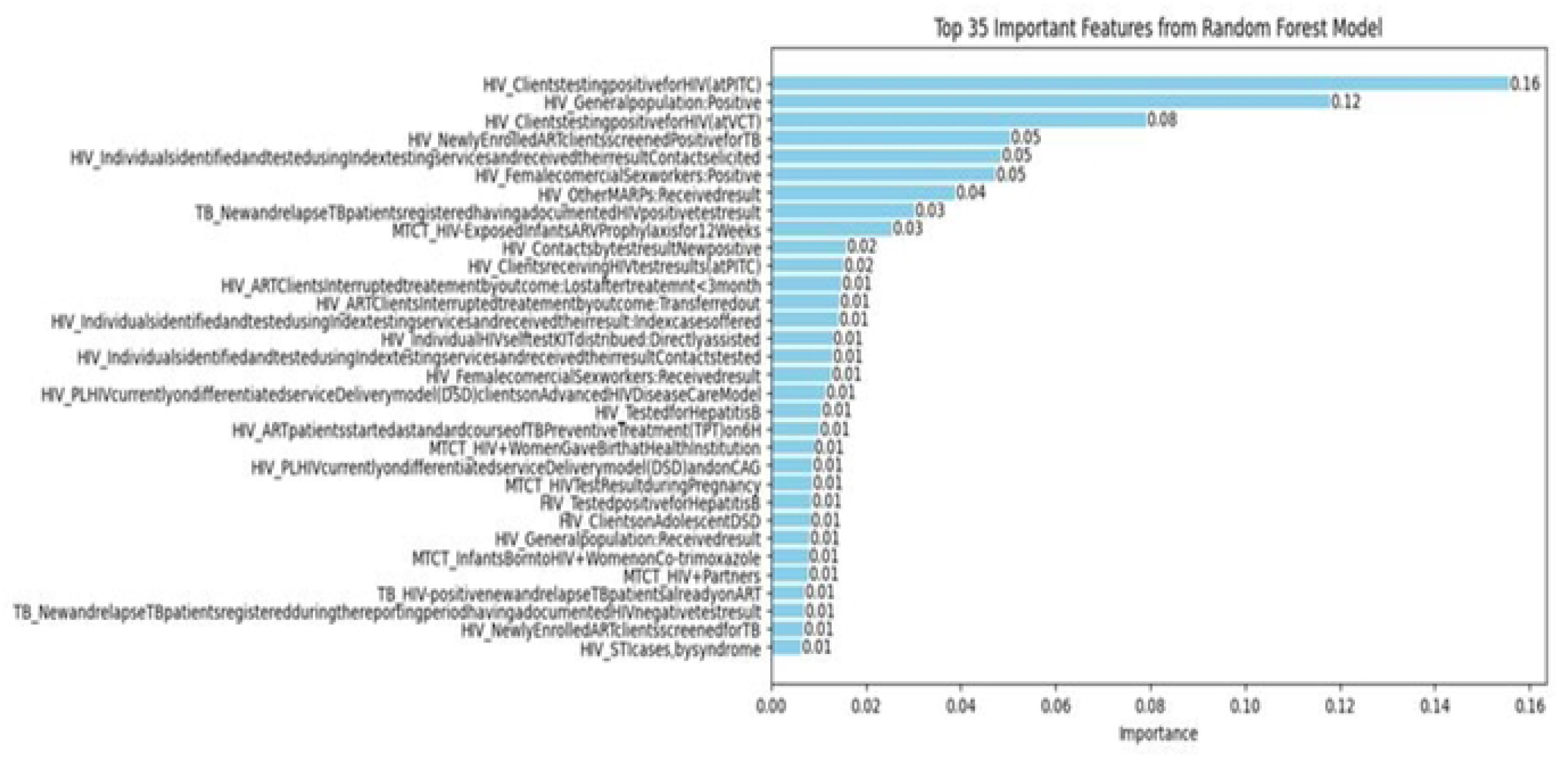
RNN model performance for HIV infection in East Gojjam zone.

##### BI directional RNN for Forecasting HIV infection in East Gojjam zone

Test MAE (original scale): 7.4618 Test MSE: 98.4083

Test RMSE: 9.9201

Test MAPE: 10.42%

**Figure 7:**
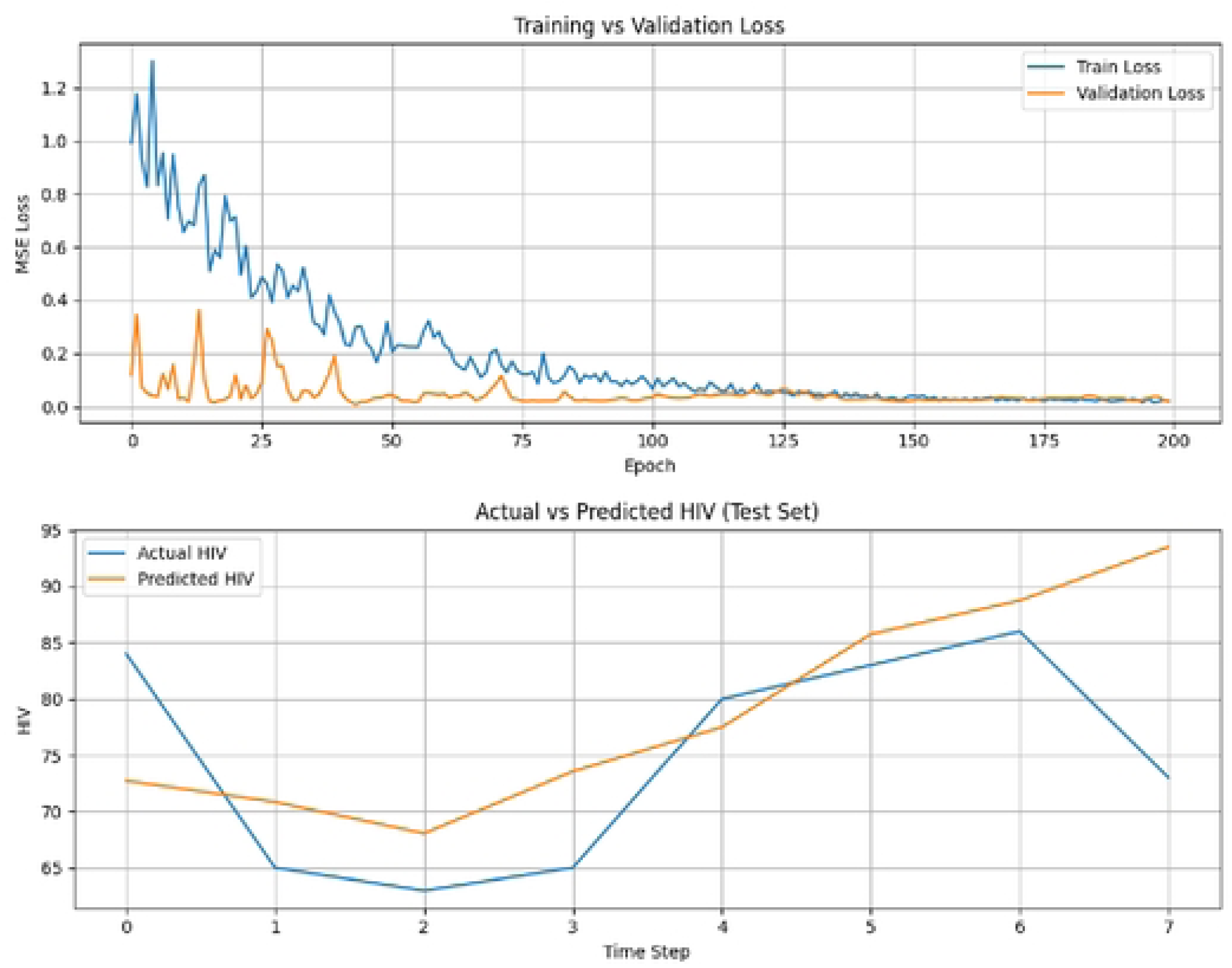
Bi-RNN model performance for HIV infection in East Gojjam zone.

##### LSTM for HIV infection in East Gojjam zone

Test MAE (original scale): 12.3919 Test MSE: 186.3182

Test RMSE: 13.6498

Test MAPE: 16.79%

**Figure 8:**
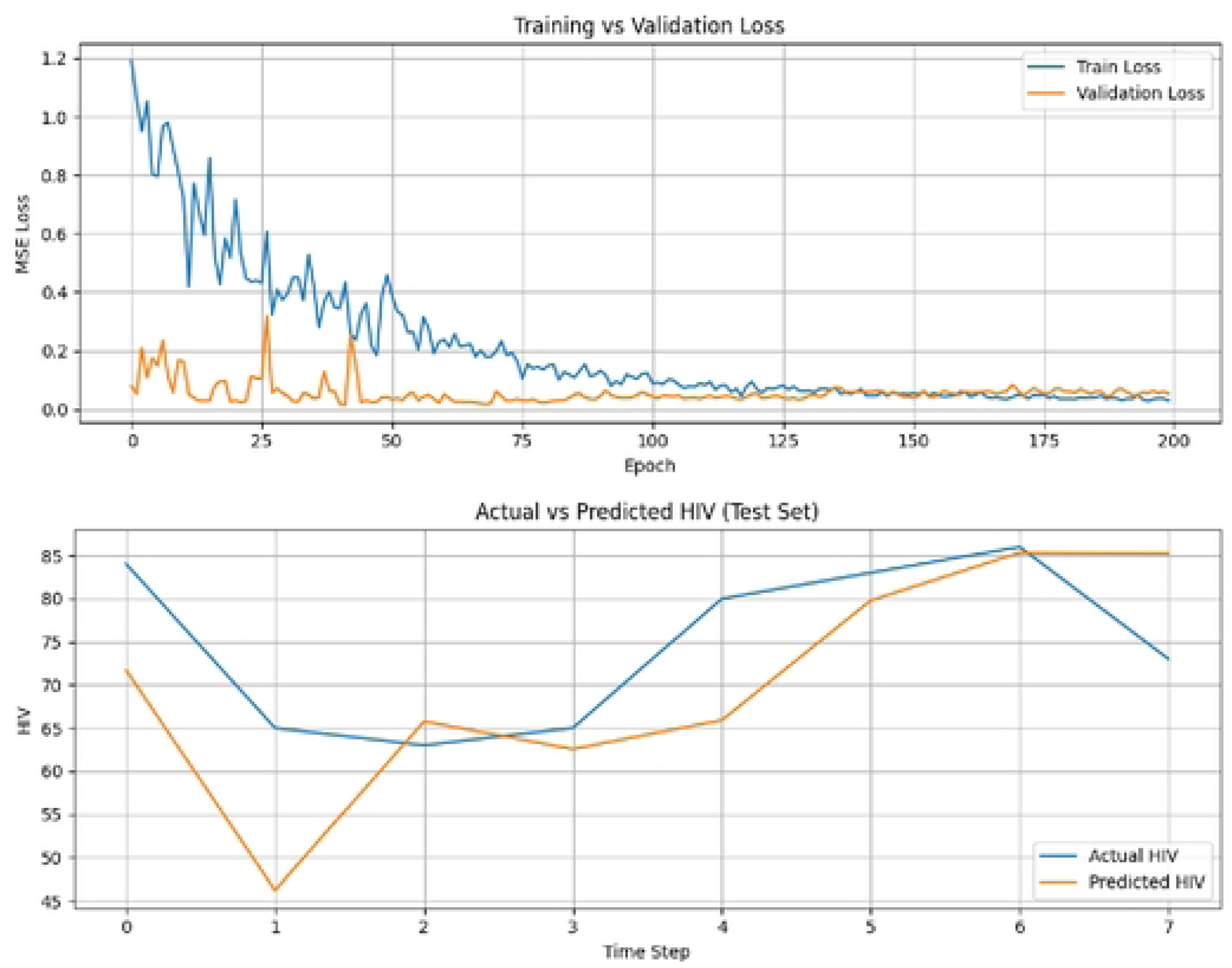
LSTM model performance in HIV infection forecasting in East Gojjam zone.

##### Bidirectional LSTM for HIV infection forecasting in East Gojjam zone

Test MAE (original scale): 6.05 Test MSE: 54.50

Test RMSE: 7.38

Test MAPE: 7.85%

**Figure 9:**
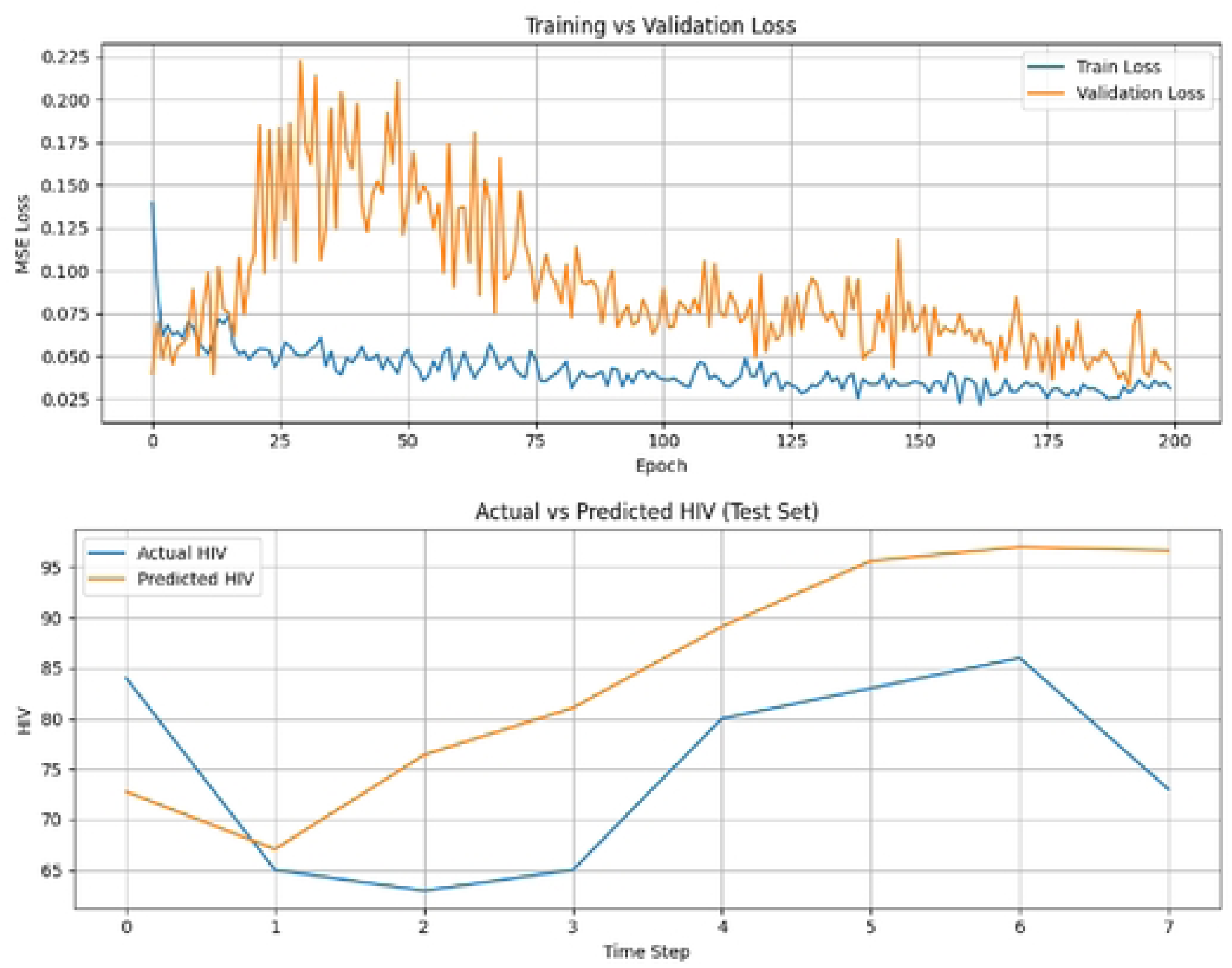
Bi-LSTM model performances in HIV infection in East Gojjam zone.

##### GRU model for HIV infection forecasting in East Gojjam zone

Test MAE (original scale): 22.4461 Test MSE: 693.9278

Test RMSE: 26.3425

Test MAPE: 29.72%

**Figure 10:**
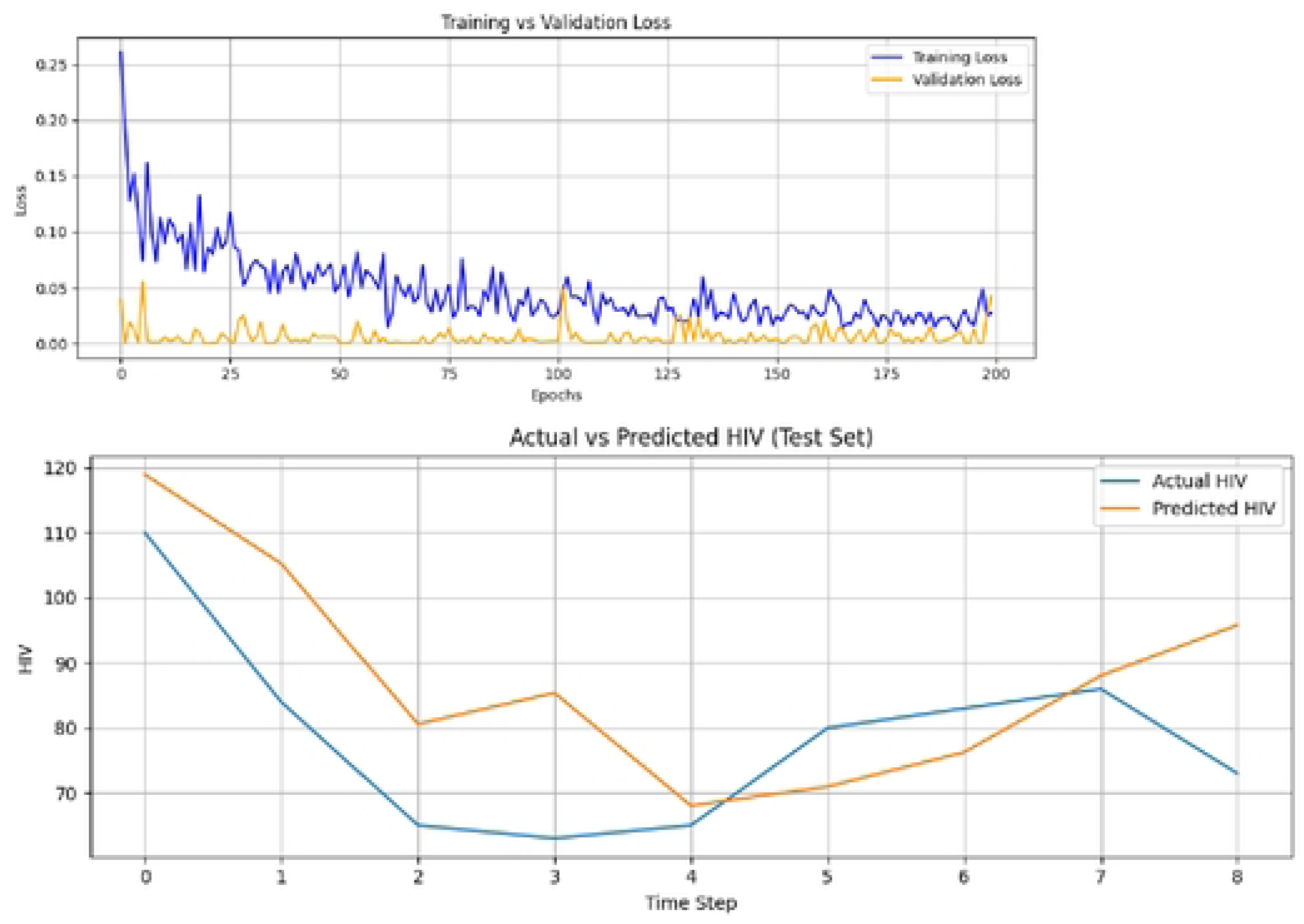
GRU model performances in HIV infection in East Gojjam zone.

##### Bidirectional GRU for HIV infection forecasting in East Gojjam zone

Test MAE (original scale): 24.2962 Test MSE: 727.8137

Test RMSE: 26.9780

Test MAPE: 32.27%

**Figure 11:**
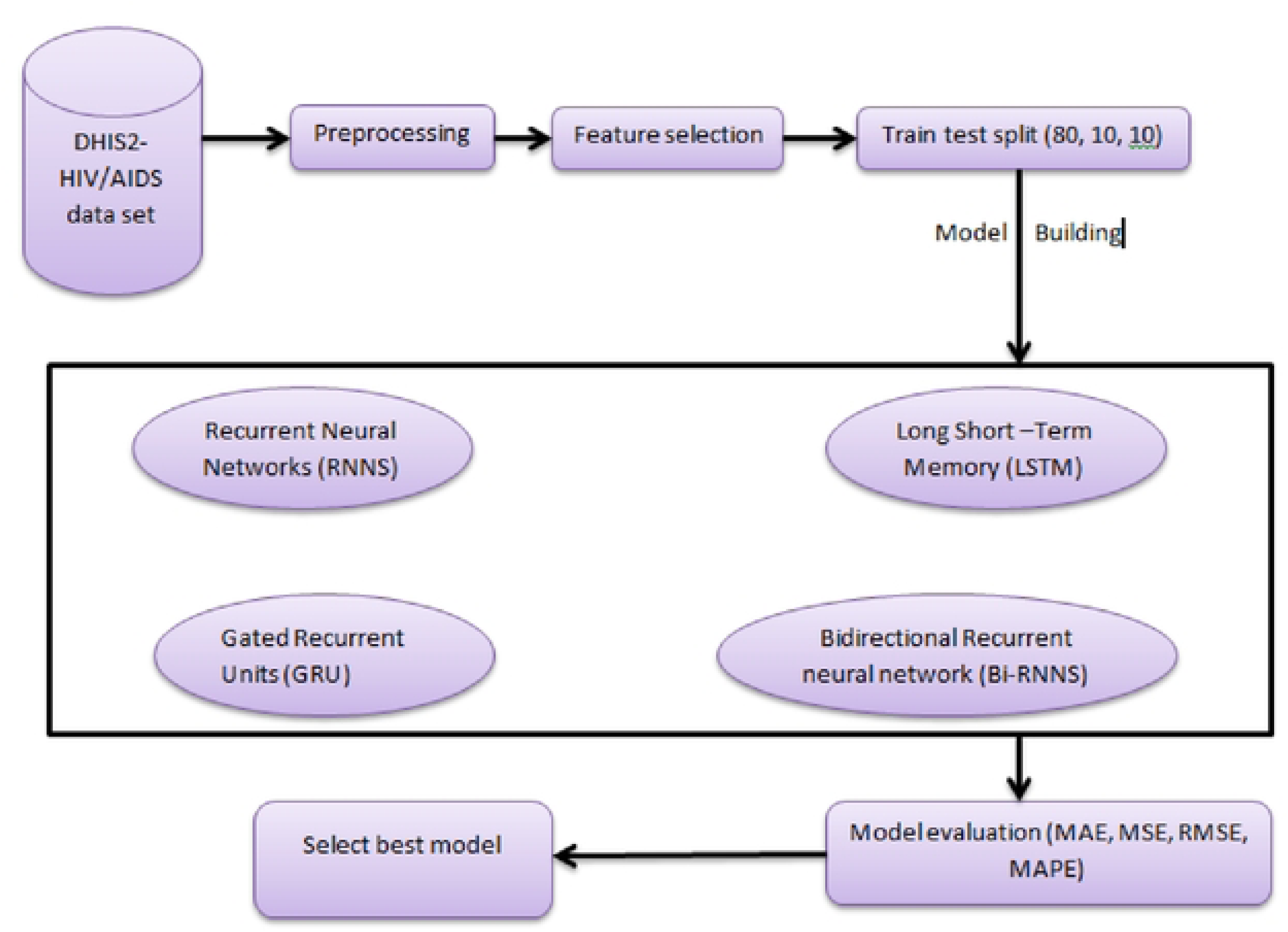
BI-GRU model performances in HIV infection in East Gojjam zone.

##### Forecasted HIV infection trends in East Gojjam zone

**Figure 12:**
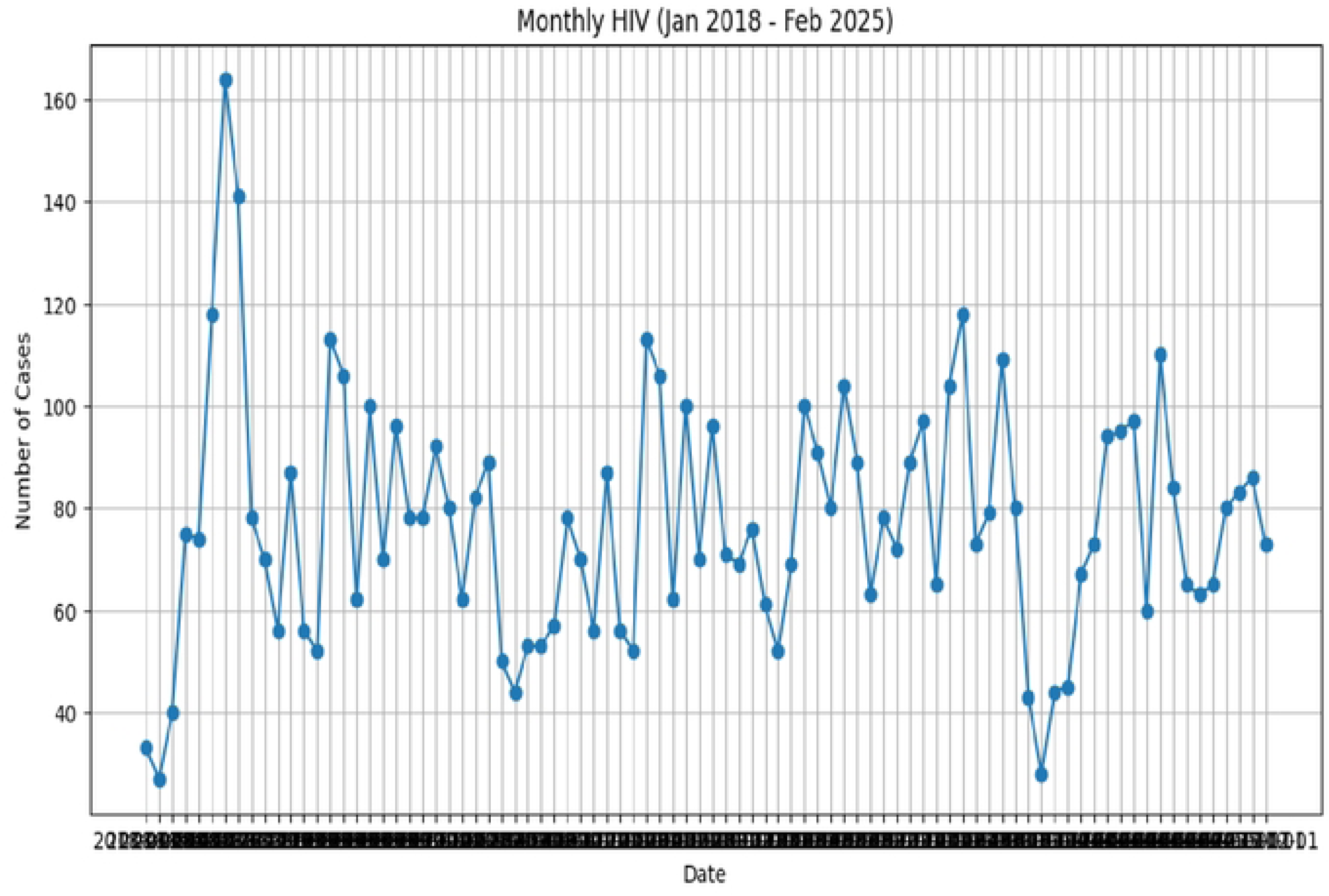
Forecasted HIV infection for next five years in east Gojjam zone using Bi LSTM.

## Discussion

The study’s hybrid machine learning and deep learning approach proved essential in identifying important HIV infection predictors and enhancing the forecast of HIV infection trends in the East Gojam Zone over the following five years. The Bidirectional LSTM had the best forecasting accuracy, while the Random Forest approach was the best feature selection model, according to the predictive findings. The forecasted rise in HIV infections over the next five years points to a severe public health concern that requires urgent attention. Random Forest was successful in identifying significant determinants of HIV infection, just like other earlier research. Other earlier research had also demonstrated how well Random Forest predicted HIV status in various contexts.

Provider-initiated counseling and testing (PICT); HIV positive clients in the general tested population; clients testing HIV positive at voluntary counseling and testing (VCT) centers; new ART clients who also screened positive for TB; and female commercial sex workers were the five most significant predictors among the numerous ones found by the Random Forest algorithm. All of these variables are essential components of standard HIV detection, prevention, and control programs and can serve as reliable predictors of new HIV incidence forecasting. The evidence found in the literature is also consistent with their relationship with HIV infection. A previous study revealed that random forest algorithms best performed in many different studies like HIV prediction using socio demographic, behavioral and biological data [55],similarly best performed in HIV drug resistance [56],In addition to this it is also well performed in virological failure in HIV/AIDS patients in studies done in University of Gondar[57]. So this shows that random forest is best model in HIV/AIDS data.

In terms of forecasting, the Bidirectional LSTM’s final performance outperforms deep learning models (RNN, LSTM, GRU, or their variants), which is consistent with findings from research on time-series modeling and infectious illness prediction. According to our findings, bi-LSTMs are particularly helpful for predicting HIV infections because they have additional capacity to capture bidirectional sequences and long-term temporal relationships.

The Bi-LSTM’s overall success in this study encourages the field to use the model for health forecasting in Ethiopia and elsewhere.[58],similarly in other studies Bi LSTM network well performed than other algorithms in Electricity forecasting [59]. The upward trend of HIV incidence in East Gojjam Zone is concerning. Despite the availability of ART, preventative programs, and testing program alternatives, the model indicates that current efforts are unlikely to reverse increases in incidence. Previous reports indicating the Amhara region continues to have persistently high HIV incidence rates corroborate this. [26, 60]. If these forecasts come true, the health care systems will be under increased pressure, and HIV/AIDS-related morbidity and mortality will rise.

These findings emphasize the necessity of targeting from a policy perspective, with preventative and testing efforts concentrating on the high-risk groups that were examined. Furthermore, it’s critical to improve ART adherence support, increase community-level HIV education, and improve PMTCT. In addition to incorporating predictive modeling into health planning, all of this will give health policy planners fast and actionable information on priorities and intervention procedures, which will ultimately result in better choices.

This research has also highlighted the drawbacks of utilizing deep learning in a low-resource context. While Bi-LSTM showed the best performance, it is unclear whether it can be adopted directly at the district level because of its computing requirements and specialized knowledge requirements. Future research should explore modified versions of a simplified version, or a hybrid model that demonstrates an acceptable performance while considering feasibility for operationalization at Ethiopian health offices.

## Conclusion

This study demonstrated the benefits of using deep learning and machine learning techniques to forecast HIV infection rates in the East Gojjam Zone. The Bidirectional LSTM achieved the most accurate forecasts (MAE = 6.05, RMSE = 7.38, MAPE = 7.85%), whereas Random Forest was found to be the best model for identifying important predictors of new infections. The forecasted upward trend in HIV infections over the course of the five years emphasizes the urgent need for HIV preventive and control measures.

The rising forecast suggests that present strategies may not be adequate to avert transmission and that certain intervention strategies are likely to create gaps. It is feasible to avoid duplication in prevention and support programs by focusing on the identified HIV high-risk populations, such as female commercial sex workers, customers of volunteer and provider-initiated testing centers, and patients who are co-infected with HIV and TB.

This study showed promise for using cutting-edge predictive algorithms that can produce useful outcomes. In general, the use of artificial intelligence to Ethiopian health surveillance systems should be taken into account. Policymakers and medical professionals can utilize predictive models to help them plan and allocate resources and target certain populations for interventions.

## Data Availability

The data will be available up on request

## Declarations

### Consent for publication

NA

### Availability of data

The data will be made available upon request.

## Author’s contribution

The first author conceptualized and designed the study, collected the data and all authors collaboratively contributed to data analysis, drafting the manuscript, and approving the final version.

## Acknowledgement

We would like to express our gratitude to Debre Markos comprehensive specialized hospital staffs that supported and contributed to this project by providing the data. Next, would like to thank Debre Markos University Department of Health Informatics for their valuable support.

## List of abbreviations

AIDS: Acquired Immunodeficiency Syndrome
AIM: AIDS Impact Model
ART: Antiretroviral therapy
BED-CEIA: Broadly Enzyme Detection–capture enzyme Immunoassay
BI-RNN: Bidirectional neural network
DL: Deep learning
GBD: Global Burdens of Disease
GRU: Gated recurrent Unit
HEI: HIV exposed infants
HIV: Human Immunodeficiency Virus
KNN: k-nearest neighborhood
LSTM: long short-term memory
ML: machine learning
MSM: Men who have sex with men
PLWH: People living with HIV/AIDS
RNN: recurrent neural network
SVM: Support vector machine
WHO: world Health Organization
XGBoost: Extreme Gradient Boosting

